# Association of plasma biomarkers with longitudinal change in age- and Alzheimer’s disease-related brain atrophy patterns

**DOI:** 10.64898/2026.09.11.26362869

**Authors:** Murat Bilgel, Ishaan Shah, Jasmine Cooper, Yang An, Keenan A. Walker, Sara G. Ho, Abhay R. Moghekar, Zhijian Yang, Guray Erus, Christos Davatzikos, Luigi Ferrucci, Susan M. Resnick

## Abstract

**INTRODUCTION:** Determining whether plasma biomarkers are preferentially associated with Alzheimer’s disease (AD)-related rather than age-related brain atrophy patterns may clarify their prognostic and diagnostic clinical use.

**METHODS:** Using data from the Baltimore Longitudinal Study of Aging (*N*=818), we examined cross-sectional plasma Aβ_42_/Aβ_40_, GFAP, NfL, p-tau181, and p-tau217 measurements obtained while participants were cognitively unimpaired (CU). During follow-up, 104 participants developed mild cognitive impairment (MCI)/dementia (74 due to AD, 24 due to non-AD, 6 unknown etiology). 2,293 longitudinal brain MRIs were used to quantify multidimensional atrophy pattern scores reflecting brain age (SPARE-BA), AD-like patterns (SPARE-AD), and five dominant dimensions of atrophy (R-indices). We investigated the associations of plasma biomarkers and atrophy pattern scores at index visit with conversion to MCI/dementia due to AD. We then examined the associations of plasma biomarkers with longitudinal change in pattern scores using linear mixed effects models.

**RESULTS:** p-tau181, Aβ_42_/Aβ_40_ (Lumipulse), and p-tau217 were associated with incident MCI/dementia due to AD but not non-AD etiologies, while GFAP was associated with incident MCI/dementia due to both AD and non-AD. All four biomarkers were associated with longitudinal SPARE-AD changes. Aβ_42_/Aβ_40_ (Quanterix and Lumipulse), p-tau181, and p-tau217 were associated with longitudinal parieto-temporal atrophy. p-tau217 was the only biomarker associated with longitudinal medial temporal lobe atrophy. We did not find associations between plasma biomarkers and SPARE-BA or R-indices capturing subcortical, diffuse cortical, or perisylvian atrophy.

**DISCUSSION:** Among CU individuals, plasma p-tau217 was associated with subsequent MCI/dementia due to AD and showed the most extensive associations with longitudinal AD-related brain atrophy.

## 1 Background

Alzheimer’s disease (AD) is the most common cause of dementia and is marked by cerebral amyloid-β (Aβ) plaque accumulation, neurofibrillary tau tangles, and neurodegeneration. These brain alterations begin years before the onset of cognitive symptoms and are evident on magnetic resonance imaging (MRI) and positron emission tomography (PET) scans [1]. Plasma protein assays allow for the indirect assessment of these brain changes. Given that they are more convenient than brain MRI and PET scans or lumbar punctures for measuring protein levels in the cerebrospinal fluid (CSF), they can be more readily adopted for use in clinical practice [2]. Numerous publications have reported on the agreement of plasma biomarkers with brain Aβ and tau tangle pathology. However, the literature on the associations of plasma biomarkers with neurodegeneration remains limited, and it is unclear whether plasma biomarkers can distinguish between age-related and AD patterns of atrophy. Examining the associations of plasma biomarkers with longitudinal AD-versus age-related brain atrophy among cognitively unimpaired (CU) individuals may help clarify their potential utility as indicators of subsequent AD-related brain atrophy.

Prior work has investigated longitudinal change in the volume or cortical thickness of individual brain regions in relation to plasma measures related to amyloid and tau proteinopathies. Tissot et al. showed that phosphorylated tau at threonine 181 (p-tau181) among CU individuals predicted longitudinal gray matter deterioration in the temporal lobe, precuneus, and anterior cingulate cortex [3]. Dark et al. reported associations between higher baseline p-tau181 and steeper gray matter volume decline in temporal regions [4]. Wang et al. found that among CU individuals with elevated brain Aβ pathology (as measured via PET imaging or CSF Aβ1-42), higher p-tau181 was associated with steeper hippocampal volume loss [5]. Plasma p-tau217 may offer superior prognostic value for predicting longitudinal cognitive decline and downstream biological changes in preclinical AD [6,7], but its association with longitudinal brain atrophy has been less extensively characterized. In a sample that included both CU individuals and individuals with subjective cognitive decline or mild cognitive impairment (MCI), Pereira et al. found an association between higher p-tau217 and steeper parietal lobe atrophy over time [8]. There are fewer studies reporting associations between plasma Aβ and atrophy in AD-related brain regions compared to the plasma p-tau literature. Among CU participants, lower plasma Aβ_42_/Aβ_40_ was associated with gray matter loss in lateral temporal regions [9]. In contrast, other studies have reported a lack of an association between plasma Aβ_42_/Aβ_40_ and hippocampal volume [4,10].

Other commonly examined plasma biomarkers in the context of AD include neurofilament light chain (NfL) and glial fibrillary acidic protein (GFAP). NfL, a marker of axonal damage, increases in preclinical AD in response to amyloid-related neuronal injury [11]. Plasma NfL has been associated with hippocampal atrophy [9,12] and volume loss in other AD-vulnerable regions among individuals with elevated amyloid-beta burden (Aβ+) [13]. GFAP, a marker of astrocyte reactivity, has also been associated with lower cortical thickness among Aβ+ individuals [14,15].

In this study, rather than focusing on individual brain regions, we examine the associations of plasma Aβ_42_/Aβ_40_, GFAP, NfL, p-tau181, and p-tau217 with longitudinal rates of change in multidimensional brain atrophy pattern scores among CU individuals. These atrophy scores capture distributed patterns of brain atrophy associated with aging and neurodegenerative processes. We address an important gap in the literature by examining whether plasma biomarkers are preferentially associated with longitudinal change in AD-related versus age-related atrophy patterns. Compared with normal aging, AD is associated with marked atrophy in medial temporal regions, particularly the entorhinal cortex and hippocampus early in the disease, with additional involvement of temporal and parietal association cortices as the disease progresses [16]. We hypothesize that plasma Aβ_42_/Aβ_40_, p-tau181, and p-tau217, which reflect AD proteinopathies, will be associated with longitudinal changes in AD-like brain atrophy, whereas GFAP and NfL, which are non-specific measures of astrocyte reactivity and neuronal damage, respectively, will be associated with changes in age-related atrophy. Determining which plasma biomarkers are preferentially associated with early longitudinal AD-like atrophy patterns could motivate subsequent studies evaluating their predictive and clinical utility.

## 2 Methods

### 2.1 Participants

Our sample consisted of 818 participants from the Baltimore Longitudinal Study of Aging (BLSA), which is a continuous enrollment cohort study of community-dwelling adults. All participants were cognitively unimpaired at the time of plasma collection. We used plasma biomarkers at the index visit only, which is defined for each individual as the earliest visit with MRI and plasma biomarkers. Participants underwent T1-weighted MRI scans to quantify brain atrophy, with the frequency of imaging increasing from approximately every 4 years at age < 60 to every other year at age 60 to 79 and every year at age ≥ 80. MRIs concurrent with and following the index visit, as well as those preceding the index visit by up to 5 years, were included in the analyses to improve the estimation of the rate of change in neurodegeneration.

Research protocols were conducted in accordance with United States federal policy for the protection of human research subjects contained in Title 45 Part 46 of the Code of Federal Regulations, approved by local institutional review boards, and all participants gave written informed consent at each visit.

### 2.2 Case conferencing and cognitive diagnosis

Clinical and selected neuropsychological data from BLSA participants are reviewed at a consensus conference if participants screen positive on the Blessed Information-Memory-Concentration Test score (i.e., score ≥ 4) [17], if their Clinical Dementia Rating (CDR) score is ≥ 0.5 using subject or informant report [18], or if concerns are raised about their cognitive status. MCI is determined using the Petersen criteria and diagnosed when (1) cognitive impairment is evident for a single domain (typically memory) or (2) cognitive impairment in multiple domains occurs without significant functional loss in activities of daily living [19]. Both cross-sectional and longitudinal neuropsychological performance, as well as the CDR, are used to determine cognitive impairment. Neuropsychological tests used in the diagnosis include Mini-Mental State Examination (MMSE) [20], Trail Making Test [21], Letter (F, A, S) and Category (animals, vegetables, fruits) Fluency [23], Boston Naming Test [24], Clock Drawing Test [25], Constructions, Calculations, and a memory impairment screen with maximum score of 13 (MIS+). The MIS+ is based on the 4-item short delay free and cued recall score from the MIS test (maximum score = 8) [26] plus the 5-item memory score from the Blessed Information-Memory-Concentration Test (maximum total score = 13). The use of MIS+ has been validated against the use of the Picture Version Free and Cued Selective Reminding Test as a memory screen for the diagnosis of MCI and dementia. Diagnoses of dementia and AD follow the Diagnostic and Statistical Manual, the third edition, revised (DSM-III-R) [27] and the National Institute of Neurological and Communication Disorders and Stroke-Alzheimer’s Disease and Related Disorders Association (NINCDS-ADRDA) criteria [28], respectively.

### 2.3 Plasma biomarkers

Plasma biomarkers were quantified using Quanterix Simoa and Fujirebio Lumipulse assays.

#### 2.3.1 Quanterix Simoa assays

Plasma Aβ_40_, Aβ_42_, GFAP, and NfL concentrations were measured using the Quanterix Simoa Neurology 4-plex-E (N4PE) and p-tau181 was measured using the Quanterix pTau-181 V2 singleplex on the Quanterix HD-X instrument. Most samples were measured in duplicate, with a small number measured in singlicate or triplicate. Singlicates were excluded from further analysis. Concentrations below the limit of detection were set to half of the limit of detection, and those above the maximum of the dynamic range were set to the maximum of the dynamic range. Using the replicates, we calculated the intra-assay coefficient of variation (CV) as the ratio of the standard deviation to the mean. Measurements with an intra-assay CV above 20% were excluded from the analysis. Aβ_40_ < 20, Aβ_42_ > 25, GFAP > 1,000, or NfL > 125 pg/mL were designated as outliers and excluded. For the samples included in this analysis, the mean intra-assay CVs for Aβ_40_, Aβ_42_, GFAP, NfL, and p-tau181 were 1.9%, 2.3%, 4.1%, 3.4%, and 6.3%, respectively. We averaged the replicates within each sample and protein.

#### 2.3.2 Fujirebio Lumipulse assays

Plasma Aβ_40_, Aβ_42_, and p-tau217 were measured using the Fujirebio Lumipulse assay on the Lumipulse G1200 instrument. Concentrations were determined via a lot-specific calibration curve and were assayed in singlicate. Quality control procedures were performed at the beginning of each test day to ensure that low and high control values were within target ranges. The limit of detection for p-tau217 was 0.030 pg/mL and the dynamic range was 0.030–10 pg/mL. The inter-assay CVs for Aβ_40_, Aβ_42_, and p-tau217 were 3.36% (high control) and 3.48% (low control), 5.95% (high control) and 5.60% (low control), and 2.02% (high control) and 4.99% (low control), respectively. In this analysis, Aβ_40_ > 1,000 or p-tau217 > 3 pg/mL were designated as outliers and excluded.

#### 2.3.3 Preprocessing

We calculated the ratio of Aβ_42_ to Aβ_40_ for the Quanterix and Fujirebio Lumipulse assays. We natural log-transformed GFAP, NfL, p-tau181, and p-tau217 to account for their skewed distributions. Variables were *z*-scored prior to statistical analysis.

We estimated glomerular filtration rate (eGFR) at each plasma visit from serum creatinine levels using the Chronic Kidney Disease-Epidemiology collaboration formula [29]. For visits without serum creatinine measurements, we imputed eGFR by carrying it forward or backward in time within person.

### 2.4 Brain magnetic resonance imaging

T1-weighted MRIs were acquired using a magnetization-prepared rapid gradient echo (MPRAGE) sequence on a 3 T Philips Achieva scanner (repetition time = 6.8 ms, echo time = 3.2 ms, flip angle = 8°, image matrix = 256 × 256, 170 slices, pixel size = 1 × 1 mm, slice thickness = 1.2 mm, sagittal acquisition).

Each scan was segmented into a set of anatomical regions of interest (ROIs) using a multi-atlas label fusion method, MUSE [30]. We calculated Spatial Pattern of Atrophy for Recognition of Early Brain Aging (SPARE-BA) and Spatial Pattern of Abnormality for Recognition of Early Alzheimer’s Disease (SPARE-AD) scores, which are individualized indices of age- and AD-related brain atrophy, respectively [31,32]. SPARE-BA is a quantitative characterization of structural age-related differences in brain anatomy through the human lifespan and is computed using a regression model trained to predict chronological age from brain atrophy patterns of cognitively normal participants [33,34]. SPARE-AD is an index derived from regional brain volumes of cognitively normal older adults and clinical AD patients to quantify atrophy patterns associated with AD. A high-dimensional pattern classifier is trained to identify imaging patterns that maximally differentiate between the two groups using a support vector machine, and applied on new data to predict a single summary score that quantifies AD-related atrophy [31]. SPARE-AD predicts conversion from normal cognition to MCI and then to dementia due to AD with high accuracy [35,36]. The SPARE-BA and SPARE-AD models were trained on a large and diverse multi-site sample from the iSTAGING data consortium, using site-harmonized volumes of segmented anatomical regions as input features. We also calculated R-indices describing five dominant dimensions of brain aging: subcortical (R1), medial temporal lobe (R2), parieto-temporal (R3), diffuse cortical (R4), and perisylvian (R5) [37]. These indices were calculated using semi-supervised representation learning via a generative adversarial network, Surreal-GAN, which captures these brain changes relative to a young and healthy reference population and distills them down to a low-dimensional representation. Higher values for SPARE scores and R-indices indicate greater atrophy according to the corresponding pattern.

### 2.5 Statistical analysis

#### 2.5.1 Conversion to MCI or dementia

We used Cox proportional hazards models to quantify the association of atrophy pattern scores and plasma biomarkers at index visit with time to conversion to MCI/dementia due to AD. For participants who developed MCI/dementia due to AD over their longitudinal follow-ups, we used their date of impairment onset (if an MCI diagnosis was made) or dementia diagnosis to calculate time to event. For remaining participants, time to event was right censored at last visit. The independent variable of interest was the continuous atrophy pattern score or plasma biomarker. To allow for comparison of the hazard ratios across models, atrophy pattern scores and plasma biomarkers were *z*-scored prior to fitting the models. We included age at index visit, sex, interaction between age and sex, race, years of education, and eGFR as covariates. We fitted the models and assessed the proportional hazards assumption using the `survival` package [38] in R version 4.3.3 [39]. Statistical significance was defined as two-tailed unadjusted *p* < .05. We did not apply multiple-comparison correction for these analyses as they were exploratory and intended to be supportive of the main analysis described below.

#### 2.5.2 Longitudinal change in atrophy pattern scores

We first assessed whether individuals with subsequent impairment exhibited differences in longitudinal rates of change in atrophy pattern scores (SPARE-BA, SPARE-AD, and the five R-indices) using a linear mixed effects model for each atrophy pattern score. Independent variables included age at index visit, sex, age × sex, race, years of education, eGFR at index visit, a three-level factor indicating future conversion to MCI/dementia (remained cognitively unimpaired, converted to MCI/dementia due to AD, converted to MCI/dementia not due to AD), and the interactions of each of these terms with time from index visit (as well as the main effect of time in decades). We also included an age^2^ term to capture quadratic relationships. Each continuous variable was transformed into a *z*-score. We included a random intercept and random slope for time per participant.

We then conducted our main analysis, which involved the assessment of the associations of plasma biomarkers at index visit with the longitudinal rates of change in each atrophy pattern score. These models were structured in the same manner as described above, except that we used continuous plasma biomarker values instead of the MCI/dementia conversion factor. A separate model was fitted for each plasma biomarker and atrophy pattern score, yielding a total of 42 models (6 plasma biomarkers × 7 atrophy pattern scores [2 SPARE scores + 5 R-indices]). The fixed effects of interest were the plasma biomarker at index visit, which reflects the cross-sectional association between the plasma biomarker and the atrophy pattern score, and the plasma biomarker × time interaction, which reflects the association between the plasma biomarker and the longitudinal rate of change in the atrophy pattern score.

Linear mixed effects models were fitted using the `lmerTest` package [40]. For each model, we examined the validity of statistical assumptions such as linearity, normality of residuals and random effects, homoscedasticity of residual variance, and non-collinearity of independent variables using the `performance` package [41]. Statistical significance was defined as two-tailed adjusted *p* < .05, with multiple comparisons controlled using the Benjamini-Hochberg false discovery rate (FDR) procedure across the 42 models in our main analysis.

### 2.6 Data availability

Code for performing the statistical analyses, generating the figures and tables, and compiling the manuscript is provided in an open repository (https://gitlab.com/bilgelm/spare-plasma). BLSA data are available upon request from https://www.blsa.nih.gov. Requests are reviewed by the Data Sharing Proposal Review Committee and require the establishment of a data transfer agreement between the NIA and the recipient institute.

## 3 Results

### 3.1 Descriptives

Analyses investigating associations with Quanterix N4PE measures (Aβ_42_/Aβ_40_, GFAP, and NfL) included 818 participants with 2,293 longitudinal MRI data points. The median number of longitudinal MRIs per participant was 2 (IQR 1–4), corresponding to a median follow-up duration of 3.9 (IQR 0–6.1) years. Excluding the 250 participants with a single MRI, the median follow-up duration was 4.8 (IQR 3.3–6.7) years. Participant demographics for the N4PE analyses are shown in Table 1.

**Table 1.** Participant characteristics for the Quanterix Neurology 4-plex-E sample. For continuous and categorical variables, we report the median and interquartile range or the N and percentage, respectively.

| Characteristic | N = 818 |
| --- | --- |
| Age at time of plasma collection (yr) | 73 (65, 81) |
| Male | 367 (45%) |
| Race |  |
| Black | 211 (26%) |
| White | 550 (67%) |
| Other | 57 (7.0%) |
| Education (yr) | 18 (16, 18) |
| Estimated glomerular filtration rate (mL/min/1.73 m<sup>2</sup>) | 80 (69, 90) |
| Quanterix A$\beta_{42}$/A$\beta_{40}$ | 0.054 (0.046, 0.061) |
| Quanterix GFAP (pg/mL) | 168 (117, 241) |
| Quanterix NfL (pg/mL) | 23 (16, 32) |
| Time between first and last MRI (yr) | 3.9 (0.0, 6.1) |
| Final clinical diagnosis |  |
| Cognitively unimpaired | 708 (87%) |
| MCI due to AD | 47 (5.7%) |
| MCI not due to AD | 16 (2.0%) |
| MCI, etiology unknown | 3 (0.4%) |
| Other impairment | 6 (0.7%) |
| Dementia due to AD | 27 (3.3%) |
| Dementia not due to AD | 8 (1.0%) |
| Dementia, etiology unknown | 3 (0.4%) |

Abbreviations: Aβ , amyloid-beta; AD, Alzheimer's disease; GFAP, glial fibrillary acidic protein; MCI, mild cognitive impairment; NfL, neurofilament light chain.

The Quanterix p-tau181 and Fujirebio Lumipulse Aβ_42_/Aβ_40_ and p-tau217 datasets were constructed from participants in the larger Quanterix N4PE dataset and included 526 participants (1,673 MRI data points) (Supplementary Table 1) and 428 participants (1,525 MRI data points) (Supplementary Table 2), respectively.

In a subset of participants who also underwent [^11^C]Pittsburgh compound B (PiB) PET imaging to quantify cerebral fibrillar amyloid plaque burden, we confirmed that plasma biomarkers (except for NfL) at index visit exhibited statistically significant differences between PiB negative and positive groups (Supplementary Figure 1).

Atrophy pattern scores increased with age, with SPARE-BA changing approximately linearly and SPARE-AD exhibiting a piecewise linear increase, with a change point around age 75 (Figure 1). Cross-sectionally, SPARE-BA was most strongly correlated with R3–R5, whereas SPARE-AD was most strongly correlated with R2 and R3 (Figure 2). For each atrophy pattern score, the strongest correlation among the plasma biomarkers was with NfL or GFAP. Conversely, for each plasma biomarker, the strongest correlation among the atrophy pattern scores was with SPARE-BA.

**Figure 1.**
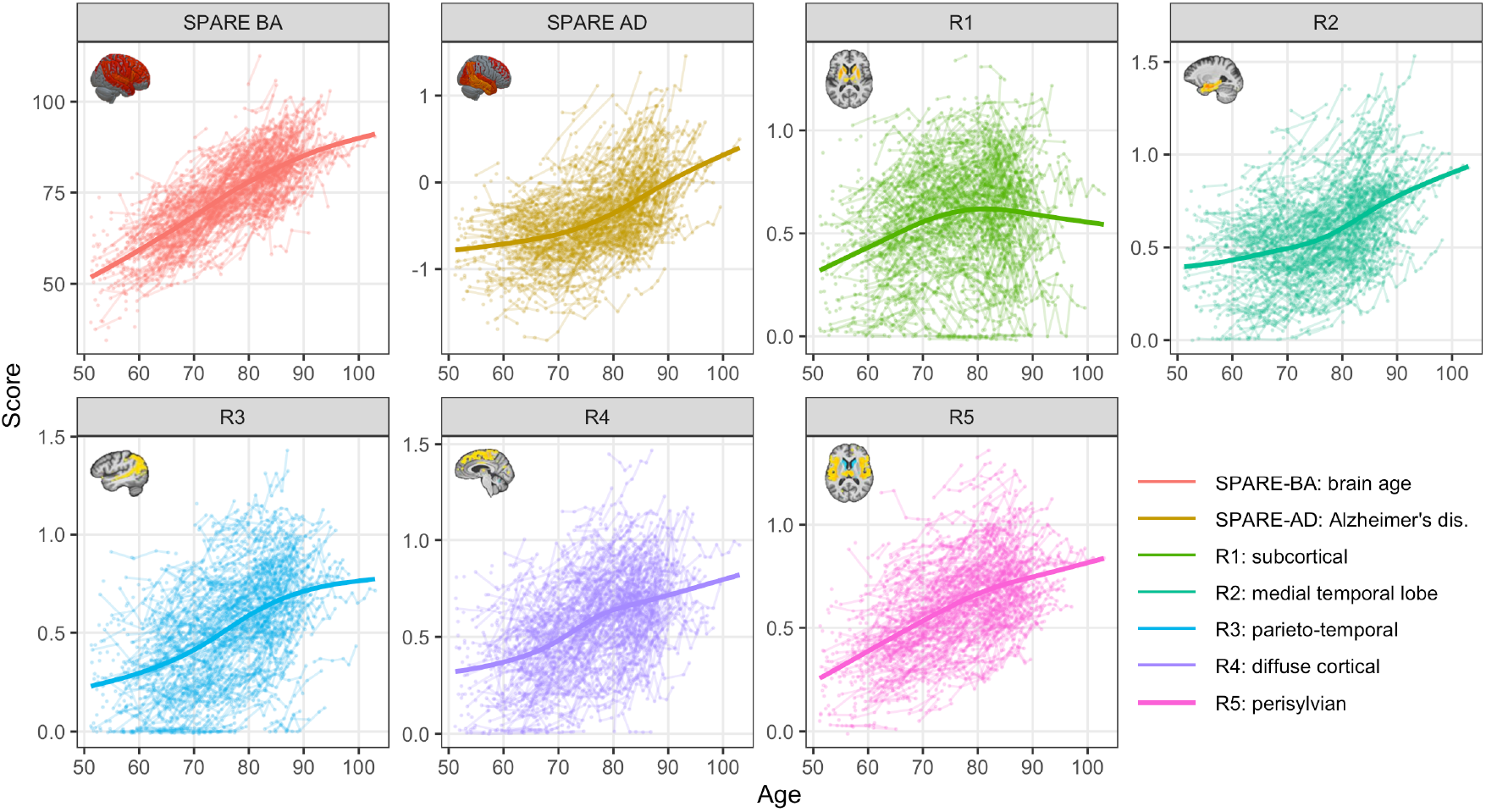
Atrophy pattern scores over time. Each line corresponds to a participant and each point corresponds to a visit. Thick curves represent the generalized additive model fits using cubic splines with shrinkage. Abbreviations: SPARE-AD, Spatial Pattern of Abnormality for Recognition of Early Alzheimer’s Disease; SPARE-BA, Spatial Pattern of Atrophy for Recognition of Early Brain Aging.

**Figure 2.**
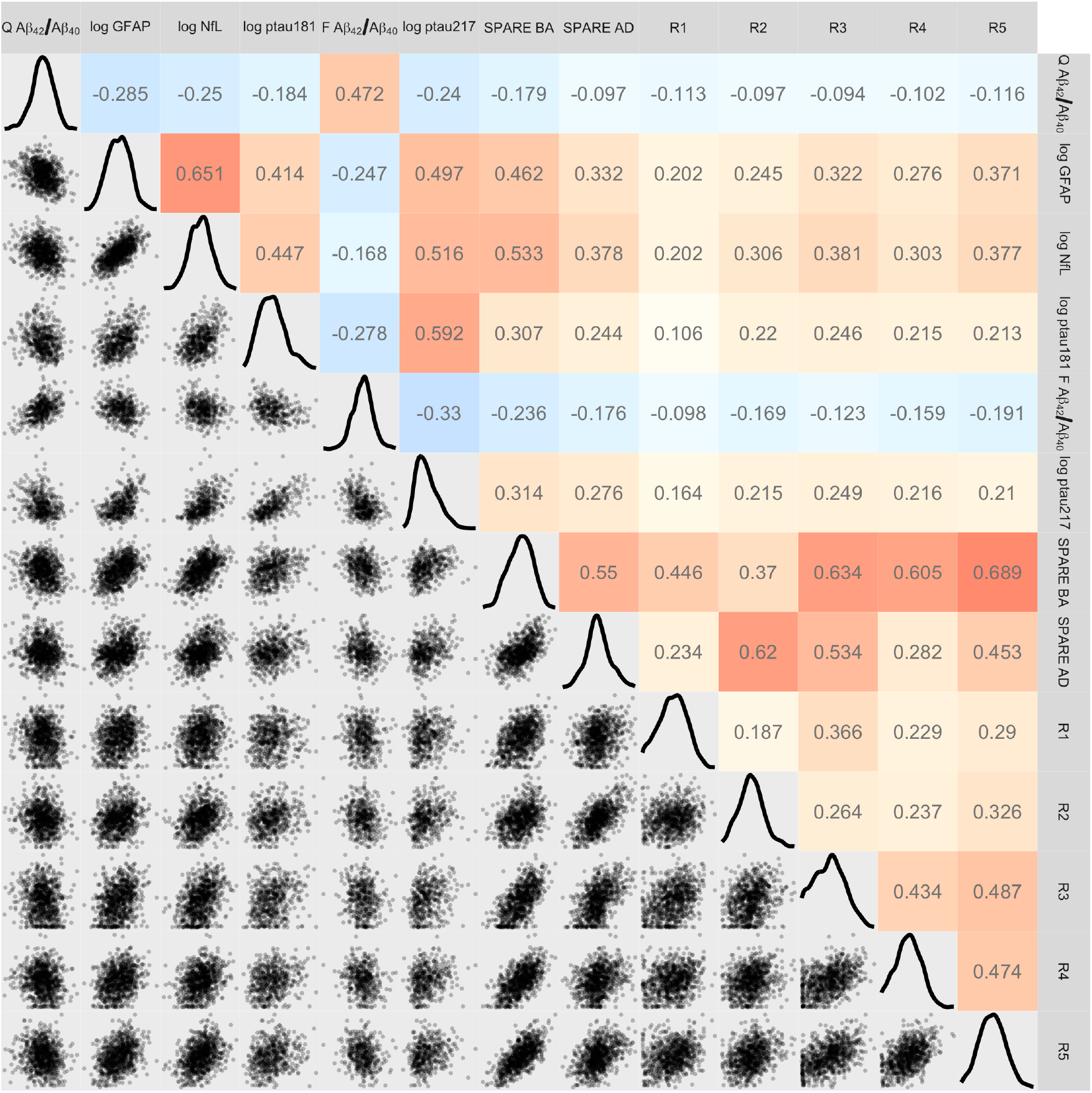
Cross-sectional correlations among plasma biomarkers and atrophy pattern scores. Lower triangular elements are pairwise scatter plots, diagonal elements are density plots, and upper triangular elements are pairwise Pearson correlations. Abbreviations: Aβ, amyloid-beta; F, Fujirebio Lumipulse; GFAP, glial fibrillary acidic protein; NfL, neurofilament light chain; Q, Quanterix Simoa; SPARE-AD, Spatial Pattern of Abnormality for Recognition of Early Alzheimer’s Disease; SPARE-BA, Spatial Pattern of Atrophy for Recognition of Early Brain Aging.

### 3.2 Multidimensional brain atrophy pattern scores and plasma biomarkers are associated with incident MCI/dementia

Of the 818 participants, 104 developed MCI or dementia during the study over a median of 3.4 (IQR 2.1–5.4) years after their plasma measurements. Of these clinical diagnoses, 74 were due to AD, 24 were due to another cause, and the etiology of the remaining 6 could not be determined. These 6 participants were included in survival analyses examining all-cause MCI/dementia but excluded from those examining MCI/dementia due to AD or due to non-AD.

Results of the adjusted Cox proportional hazards models are summarized in Figure 3. Each atrophy pattern score except for R1 had a statistically significant association with conversion to all-cause MCI/dementia (Supplementary Table 3). Higher SPARE-AD (HR = 1.9, 95% CI = [1.4, 2.5], *p* = 3.7 × 10^−6^) and R2 (HR = 1.5, 95% CI = [1.2, 1.9], *p* = 0.0023) were associated with a higher risk of conversion to MCI/dementia due to AD, whereas their associations with non-AD conversion were not statistically significant. Higher R3 (HR = 1.7, 95% CI = [1.1, 2.9], *p* = 0.031) and R4 (HR = 2.5, 95% CI = [1.5, 4], *p* = 0.00022) were associated with a higher risk of conversion to MCI/dementia due to non-AD, whereas their associations with AD conversion were not statistically significant. We also found statistically significant associations between all plasma biomarkers except Quanterix Aβ_42_/Aβ_40_ and conversion to all-cause MCI/dementia; the association for Quanterix Aβ_42_/Aβ_40_ was trend-level (*p* = 0.052) (Supplementary Table 4). Higher log GFAP *z*-score (HR = 1.5, 95% CI = [1.1, 2], *p* = 0.016), log p-tau181 *z*-score (HR = 1.7, 95% CI = [1.2, 2.4], *p* = 0.0022), log p-tau217 *z*-score (HR = 1.8, 95% CI = [1.3, 2.5], *p* = 0.00034), and lower Lumipulse Aβ_42_/Aβ_40_ *z*-score (HR = 0.6, 95% CI = [0.43, 0.84], *p* = 0.0031) were associated with a higher risk of conversion to MCI/dementia due to AD. Higher log GFAP *z*-score was also associated with a higher risk of conversion to MCI/dementia due to non-AD (HR = 2.2, 95% CI = [1.4, 3.6], *p* = 0.0013).

**Figure 3.**
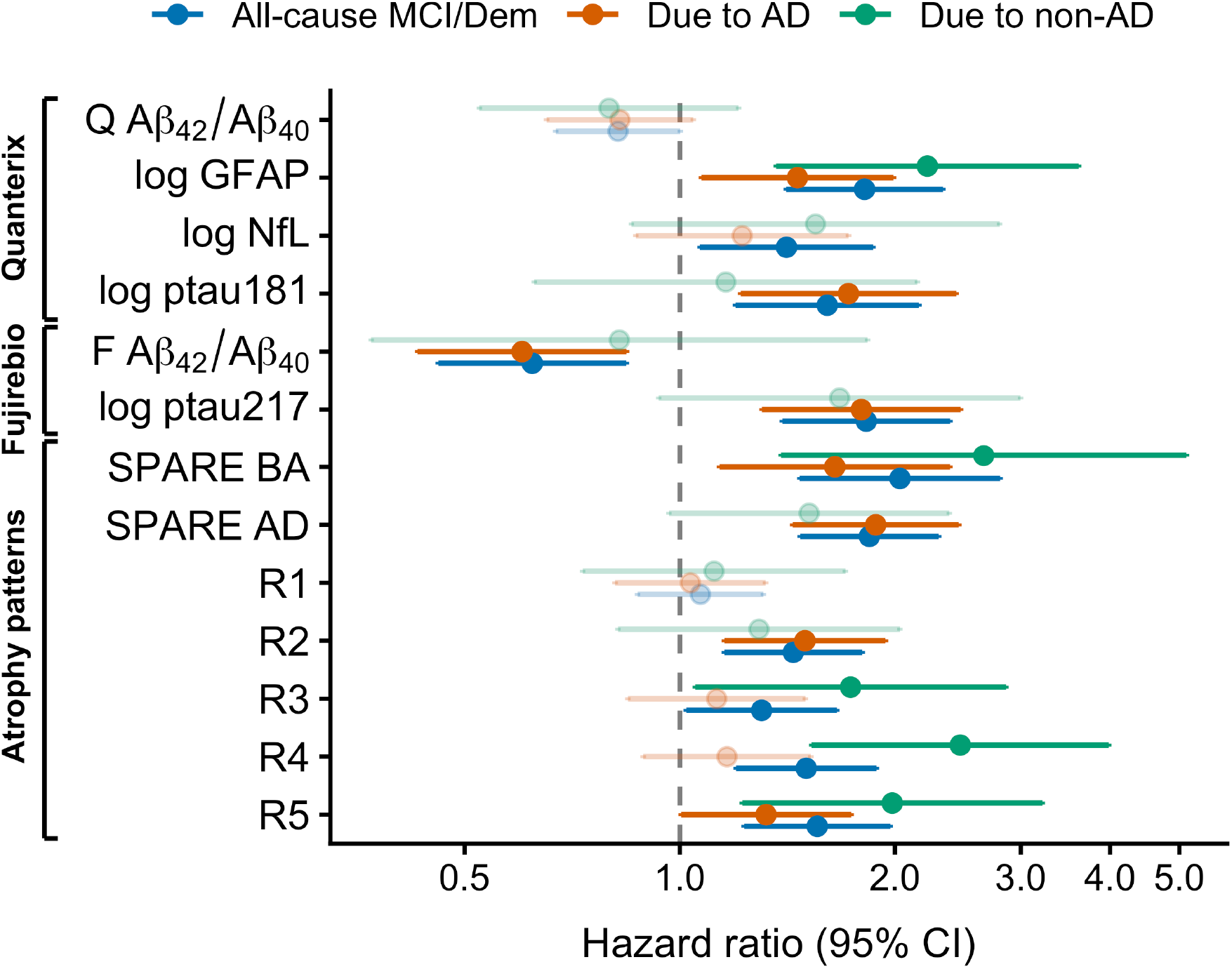
Cox proportional hazards model results. The forest plot displays the hazard ratios for z-scored plasma biomarkers and atrophy pattern scores at index visit for conversion to all-cause MCI/dementia (blue), MCI/dementia due to AD (orange), and due to non-AD (teal). Abbreviations: Aβ, amyloid-beta; CI, confidence interval; F, Fujirebio Lumipulse; GFAP, glial fibrillary acidic protein; NfL, neurofilament light chain; Q, Quanterix Simoa; SPARE-AD, Spatial Pattern of Abnormality for Recognition of Early Alzheimer’s Disease; SPARE-BA, Spatial Pattern of Atrophy for Recognition of Early Brain Aging.

In demographics- and eGFR-adjusted linear mixed effects models examining longitudinal atrophy pattern scores as outcomes, compared to participants who remained cognitively unimpaired, those who later converted to MCI/dementia due to AD had greater SPARE-BA (estimate = 2, SE = 0.91, *p* = 0.029), SPARE-AD (estimate = 0.2, SE = 0.049, *p* = 5.6 × 10^−5^), R2 (estimate = 0.068, SE = 0.028, *p* = 0.015), and R5 (estimate = 0.054, SE = 0.025, *p* = 0.030) at index visit. They also exhibited steeper longitudinal increases in SPARE-AD (estimate = 0.21, SE = 0.042, *p* = 1.3 × 10^−6^) and R2 (estimate = 0.052, SE = 0.021, *p* = 0.014) (Supplementary Figure 2). Compared to participants who remained cognitively unimpaired, those who later converted to MCI/dementia *not* due to AD had greater SPARE-BA (estimate = 3.2, SE = 1.5, *p* = 0.029), SPARE-AD (estimate = 0.16, SE = 0.079, *p* = 0.044), R4 (estimate = 0.15, SE = 0.046, *p* = 0.0014), and R5 (estimate = 0.097, SE = 0.041, *p* = 0.017) at index visit. They also exhibited steeper longitudinal increases in SPARE-AD (estimate = 0.14, SE = 0.069, *p* = 0.037) and R5 (estimate = 0.067, SE = 0.023, *p* = 0.0033). These results confirmed that there are detectable differences, both cross-sectionally and longitudinally, in brain atrophy pattern scores between CU and subsequently impaired groups.

Both the Cox proportional hazards and linear mixed effects models highlighted R2 (medial temporal lobe atrophy) as a measure particularly reflective of neurodegeneration patterns related to AD but not non-AD: higher R2 at index visit was associated with a higher risk of incident MCI/dementia due to AD, and R2 increased faster among those who subsequently developed MCI/dementia due to AD, while corresponding statistically significant associations were not observed for non-AD etiologies.

### 3.3 Plasma biomarkers are associated with multidimensional patterns of brain atrophy

We did not find any statistically significant cross-sectional associations between plasma biomarkers and atrophy pattern scores that survived multiple-comparison correction. Longitudinal associations between plasma biomarkers and atrophy pattern scores in adjusted linear mixed effects models are presented in Table 2. Higher log GFAP (estimate = 0.047, SE = 0.016, *p*FDR = 0.022), higher log p-tau181 (estimate = 0.057, SE = 0.017, *p*FDR = 0.0044), lower Lumipulse Aβ_42_/Aβ_40_ (estimate = −0.049, SE = 0.016, *p*FDR = 0.0096), and higher log p-tau217 (estimate = 0.097, SE = 0.017, *p*FDR = 1.3 × 10^−6^) at index visit were associated with steeper longitudinal increases in SPARE-AD (Figure 4). Lower Quanterix Aβ_42_/Aβ_40_ (estimate = −0.021, SE = 0.0062, *p*FDR = 0.0051) and Lumipulse Aβ_42_/Aβ_40_ (estimate = −0.027, SE = 0.007, *p*FDR = 0.0014), as well as higher log p-tau181 (estimate = 0.031, SE = 0.0073, *p*FDR = 0.00047) and log p-tau217 (estimate = 0.031, SE = 0.008, *p*FDR = 0.0013) were associated with steeper longitudinal increases in R3 (parieto-temporal atrophy) (Figure 4). Only higher log p-tau217 was associated with steeper increases in R2 (medial temporal lobe atrophy) (estimate = 0.031, SE = 0.0089, *p*FDR = 0.0044).

**Figure 4.**
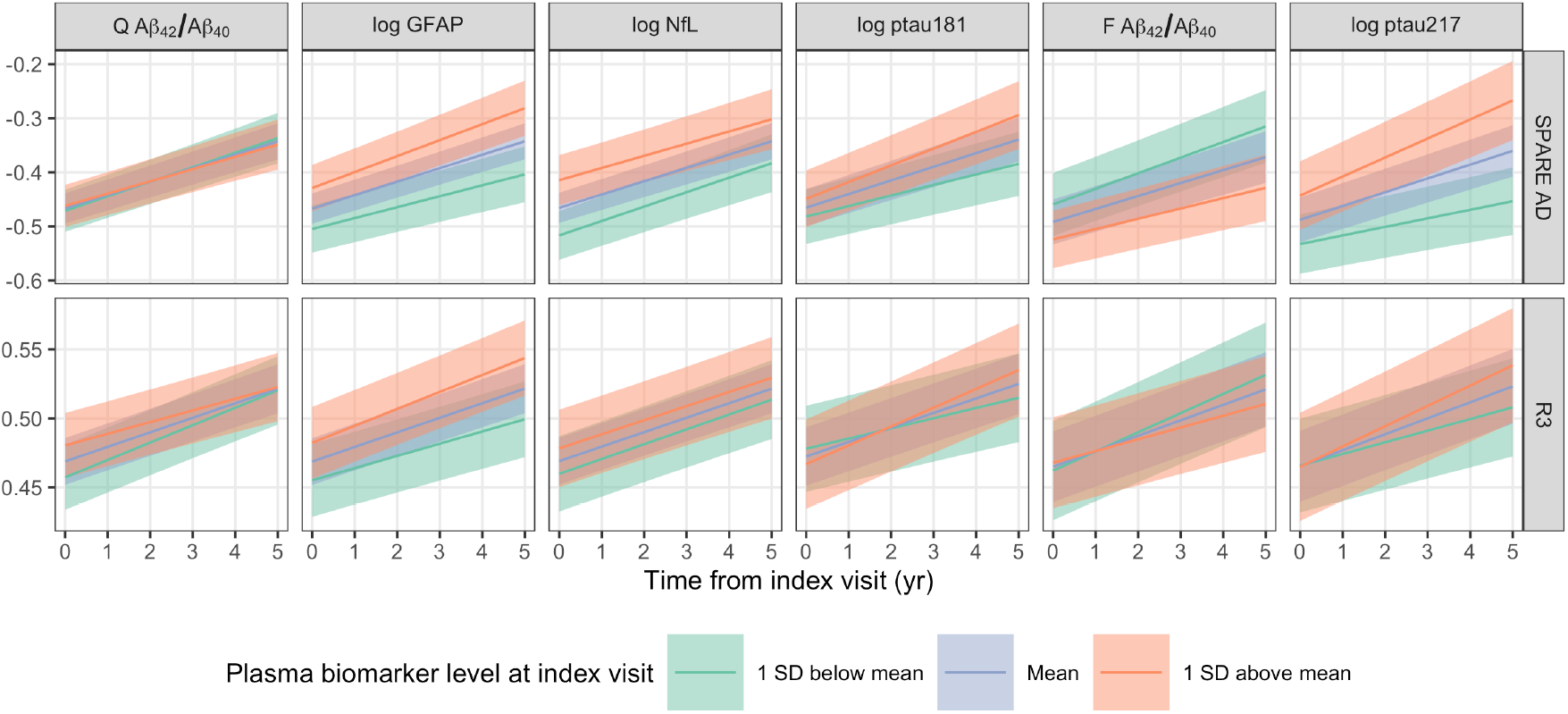
Atrophy scores by baseline plasma biomarker levels. The plasma measure used for defining the groups (indicated by the colors) in each panel is indicated at the top. Top row: Lower Lumipulse Aβ_42_/Aβ_40_ and higher GFAP, p-tau181, and p-tau217 at index visit were associated with steeper longitudinal increases in SPARE-AD. Bottom row: Lower Quanterix Aβ_42_/Aβ_40_ and Lumipulse Aβ_42_/Aβ_40_ as well as higher p-tau181 and p-tau217 were associated with steeper longitudinal increases in R3 (parieto-temporal atrophy). Abbreviations: Aβ, amyloid-beta; F, Fujirebio Lumipulse; GFAP, glial fibrillary acidic protein; NfL, neurofilament light chain; Q, Quanterix Simoa; SD, standard deviation.

**Table 2.** Results of linear mixed effects models examining the association of plasma biomarkers with longitudinal rates of change in atrophy pattern scores. Each cell corresponds to a separate model. The table shows the estimates for the biomarker × time term, which reflects the difference in the longitudinal rate of change (per decade) in atrophy pattern score with each unit standard deviation difference in the plasma biomarker at index visit. p-values are adjusted for multiple comparisons using the Benjamini-Hochberg false discovery rate (FDR) procedure.

| Outcome | Quanterix Simoa |  |  |  | Fujirebio Lumipulse |  |
| --- | --- | --- | --- | --- | --- | --- |
|  | Aβ<sub>42</sub>/Aβ<sub>40</sub> | log GFAP | log NfL | log p-tau181 | Aβ<sub>42</sub>/Aβ<sub>40</sub> | log p-tau217 |
| SPARE-BA rate | -0.272 (SE = 0.291)<br>pFDR = 0.6141 | 0.801 (SE = 0.335)<br>pFDR = 0.0628 | 0.124 (SE = 0.386)<br>pFDR = 0.8548 | 0.833 (SE = 0.347)<br>pFDR = 0.0628 | -0.620 (SE = 0.332)<br>pFDR = 0.1734 | 0.488 (SE = 0.382)<br>pFDR = 0.437 |
| SPARE-AD rate | -0.021 (SE = 0.014)<br>pFDR = 0.3417 | **0.047\* (SE = 0.016)**<br>**pFDR = 0.0218** | -0.021 (SE = 0.019)<br>pFDR = 0.5108 | **0.057\*\* (SE = 0.017)**<br>**pFDR = 0.0044** | **-0.049\*\* (SE = 0.016)**<br>**pFDR = 0.0096** | **0.097\*\*\* (SE = 0.017)**<br>**pFDR < 0.0001** |
| R1 rate | 0.003 (SE = 0.007)<br>pFDR = 0.8548 | 0.009 (SE = 0.008)<br>pFDR = 0.5108 | 0.019 (SE = 0.009)<br>pFDR = 0.1152 | -0.006 (SE = 0.008)<br>pFDR = 0.7011 | -0.000 (SE = 0.008)<br>pFDR = 0.9612 | 0.003 (SE = 0.009)<br>pFDR = 0.8548 |
| R2 rate | 0.001 (SE = 0.007)<br>pFDR = 0.9569 | 0.014 (SE = 0.008)<br>pFDR = 0.2146 | 0.014 (SE = 0.009)<br>pFDR = 0.3417 | 0.005 (SE = 0.008)<br>pFDR = 0.7989 | 0.000 (SE = 0.008)<br>pFDR = 0.9612 | **0.031\*\* (SE = 0.009)**<br>**pFDR = 0.0044** |
| R3 rate | **-0.021\*\* (SE = 0.006)**<br>**pFDR = 0.0051** | 0.017 (SE = 0.007)<br>pFDR = 0.0628 | -0.003 (SE = 0.008)<br>pFDR = 0.8548 | **0.031\*\*\* (SE = 0.007)**<br>**pFDR = 0.0005** | **-0.027\*\* (SE = 0.007)**<br>**pFDR = 0.0014** | **0.031\*\* (SE = 0.008)**<br>**pFDR = 0.0013** |
| R4 rate | -0.007 (SE = 0.008)<br>pFDR = 0.6388 | 0.011 (SE = 0.009)<br>pFDR = 0.437 | -0.001 (SE = 0.010)<br>pFDR = 0.9569 | 0.022 (SE = 0.009)<br>pFDR = 0.0683 | -0.004 (SE = 0.009)<br>pFDR = 0.8548 | 0.003 (SE = 0.010)<br>pFDR = 0.8548 |
| R5 rate | 0.003 (SE = 0.005)<br>pFDR = 0.7913 | 0.006 (SE = 0.005)<br>pFDR = 0.5108 | -0.001 (SE = 0.006)<br>pFDR = 0.9519 | 0.005 (SE = 0.006)<br>pFDR = 0.6625 | 0.003 (SE = 0.005)<br>pFDR = 0.7989 | 0.002 (SE = 0.006)<br>pFDR = 0.8548 |

Abbreviations: Aβ, amyloid-beta; GFAP, glial fibrillary acidic protein; NfL, neurofilament light chain; p-tau181, tau phosphorylated at threonine 181; p-tau217, tau phosphorylated at threonine 217; pFDR, false discovery rate-corrected p-value; SE, standard error; SPARE-AD, Spatial Pattern of Abnormality for Recognition of Early Alzheimer's Disease; SPARE-BA, Spatial Pattern of Atrophy for Recognition of Early Brain Aging. \* pFDR < 0.05, \*\* pFDR < 0.01, \*\*\* pFDR < 0.001.

## 4 Discussion

We investigated the associations of plasma biomarkers with spatial patterns of brain atrophy among older adults who were cognitively unimpaired at the time of plasma collection. Lower Aβ_42_/Aβ_40_ (Fujirebio Lumipulse), higher GFAP, p-tau181, and p-tau217 were associated with steeper longitudinal change in AD-related brain atrophy as indicated by SPARE-AD, whereas Aβ_42_/Aβ_40_, p-tau181, and p-tau217 were associated with the longitudinal rate of parieto-temporal atrophy (R3), and p-tau217 was additionally associated with the rate of medial temporal lobe atrophy (R2). These results support our initial hypothesis regarding the association between AD-like brain atrophy and plasma biomarkers that reflect AD proteinopathies, namely, Aβ_42_/Aβ_40_, p-tau181, and p-tau217, but we did not find evidence to support our hypothesized association between age-related atrophy and GFAP or NfL in models adjusted for demographics and kidney function. The preferential associations of Lumipulse Aβ_42_/Aβ_40_, p-tau181, and p-tau217 with AD-related atrophy were complemented by their associations with greater risk of conversion to MCI/dementia due to AD. Among these biomarkers, p-tau217 showed the most extensive associations across AD-related atrophy patterns and the largest effect estimate for subsequent AD-related MCI/dementia. Associations with non-AD MCI/dementia were not statistically significant, although the small number of non-AD cases limits conclusions regarding disease specificity. Overall, our results suggest that these plasma measures, p-tau217 in particular, may be useful indicators of longitudinal change in AD-related brain atrophy patterns.

Phosphorylated tau plasma biomarkers are emerging as promising indicators of AD-related brain changes. Moscoso et al. reported associations between baseline plasma p-tau181 and longitudinal atrophy in temporoparietal regions among cognitively unimpaired individuals [13], consistent with our finding that p-tau181 was associated with the rate of change in R3. Our findings agree with a growing body of literature identifying p-tau217 as a highly specific indicator of AD pathophysiology and dementia due to AD [42–45] and extend the existing literature by demonstrating the preferential association of p-tau217 with longitudinal AD-related brain atrophy patterns.

In contrast, in agreement with prior research from the BLSA examining regional brain volumes [4], we found no evidence that baseline NfL was associated with longitudinal brain atrophy after adjustment for demographics and kidney function, while GFAP was less specific to AD-related outcomes. GFAP was associated with both AD and non-AD etiologies in survival analyses and exhibited a trend towards an association with brain age as indicated by SPARE-BA. Our results are in concordance with prior reports of NfL’s poor performance as a diagnostic biomarker for AD [46]. However, NfL has been proposed as a promising biomarker for monitoring disease progression as it may reflect the degree of loss of brain structural integrity [46]. Of note, greater longitudinal increases in plasma NfL have been associated with greater hippocampal and entorhinal atrophy among cognitively unimpaired individuals [47], suggesting potential utility of longitudinal NfL measurements for preclinical AD monitoring. Further research using longitudinal plasma measures will better characterize this possibility.

Quanterix and Fujirebio Lumipulse Aβ_42_/Aβ_40_ measures were only moderately correlated, which may reflect differences between the assay technologies (Simoa vs. CLEIA). Despite the smaller Lumipulse sample, we found statistically significant associations with Lumipulse Aβ_42_/Aβ_40_ that were not statistically significant for Quanterix Aβ_42_/Aβ_40_. We did not find any statistically significant associations between Quanterix Aβ_42_/Aβ_40_ and clinical diagnosis, whereas the Lumipulse measure was associated with all-cause MCI/dementia and MCI/dementia due to AD. While both measures were associated with the rate of change in parieto-temporal atrophy (R3), only Lumipulse Aβ_42_/Aβ_40_ was associated with the longitudinal rate of change in SPARE-AD. The more widespread AD-related associations we observed for the Lumipulse assay are consistent with its higher accuracy in detecting amyloid positivity on brain PET imaging compared to Quanterix [48].

Prior research from our group examining regional brain volumes rather than atrophy pattern scores demonstrated an association between higher baseline p-tau181 and steeper declines in total gray matter volume, temporal gyri, and the amygdala, while no longitudinal brain atrophy associations were reported with Quanterix Aβ_42_/Aβ_40_ [4]. In our analyses using atrophy pattern scores, we found that both p-tau181 and Quanterix Aβ_42_/Aβ_40_ were associated with longitudinal parieto-temporal atrophy. The difference in findings may reflect the greater sensitivity of atrophy pattern scores to distributed longitudinal changes through their incorporation of multiple brain regions.

Our study has limitations. Our sample size limits our ability to reliably detect associations with small effect sizes. The BLSA is not representative of the general population, limiting the generalizability of our findings. We were unable to examine longitudinal change in plasma biomarkers due to the limited follow-up duration for plasma measurements. The relatively small number of incident non-AD cases also limited our ability to characterize biomarker associations with specific non-AD etiologies and to draw conclusions regarding the specificity of biomarker associations for AD versus non-AD outcomes.

Our study has several important strengths. The median follow-up duration for our MRI measures, 3.9 years, is longer than in most existing plasma biomarker studies investigating longitudinal associations. SPARE scores and R-indices take advantage of spatial patterns of atrophy, allowing us to capture distributed brain changes rather than focusing on individual brain regions, while mitigating the problem of multiple comparisons.

In conclusion, our results highlight plasma p-tau217 as a promising biomarker associated with longitudinal AD-related brain atrophy and subsequent MCI or dementia due to AD among cognitively unimpaired individuals.

## Data Availability

BLSA data are available upon request from https://www.blsa.nih.gov. Requests are reviewed by the Data Sharing Proposal Review Committee and require the establishment of a data transfer agreement between the NIA and the recipient institute.

## Acknowledgments

We thank the BLSA participants and staff for their dedication to these studies.

This research was supported by the Intramural Research Program of the National Institutes of Health (NIH). The contributions of the NIH authors are considered Works of the United States Government. The findings and conclusions presented in this paper are those of the authors and do not necessarily reflect the views of the NIH or the U.S. Department of Health and Human Services.

## Supplementary Material

**Supplementary Table 1.** Participant characteristics for the Quanterix p-tau181 sample. For continuous and categorical variables, we report the median and interquartile range or the N and percentage, respectively.

| Characteristic | N = 526 |
| --- | --- |
| Age at time of plasma collection (yr) | 74 (65, 81) |
| Male | 236 (45%) |
| Race |  |
| Black | 147 (28%) |
| White | 341 (65%) |
| Other | 38 (7.2%) |
| Education (yr) | 18 (16, 18) |
| Estimated glomerular filtration rate (mL/min/1.73 m<sup>2</sup>) | 79 (68, 90) |
| Quanterix Aβ<sub>42</sub>/Aβ<sub>40</sub> | 0.054 (0.046, 0.061) |
| Quanterix GFAP (pg/mL) | 174 (118, 245) |
| Quanterix NfL (pg/mL) | 22 (16, 32) |
| Quanterix p-tau181 (pg/mL) | 2.5 (1.8, 3.4) |
| Time between first and last MRI (yr) | 4.3 (2.3, 6.4) |
| Final clinical diagnosis |  |
| Cognitively unimpaired | 458 (87%) |
| MCI due to AD | 35 (6.7%) |
| MCI not due to AD | 8 (1.5%) |
| MCI, etiology unknown | 1 (0.2%) |
| Other impairment | 5 (1.0%) |
| Dementia due to AD | 12 (2.3%) |
| Dementia not due to AD | 6 (1.1%) |
| Dementia, etiology unknown | 1 (0.2%) |

Abbreviations: Aβ , amyloid-beta; GFAP, glial fibrillary acidic protein; NfL, neurofilament light chain; p-tau181, tau phosphorylated at threonine 181.

**Supplementary Table 2.**
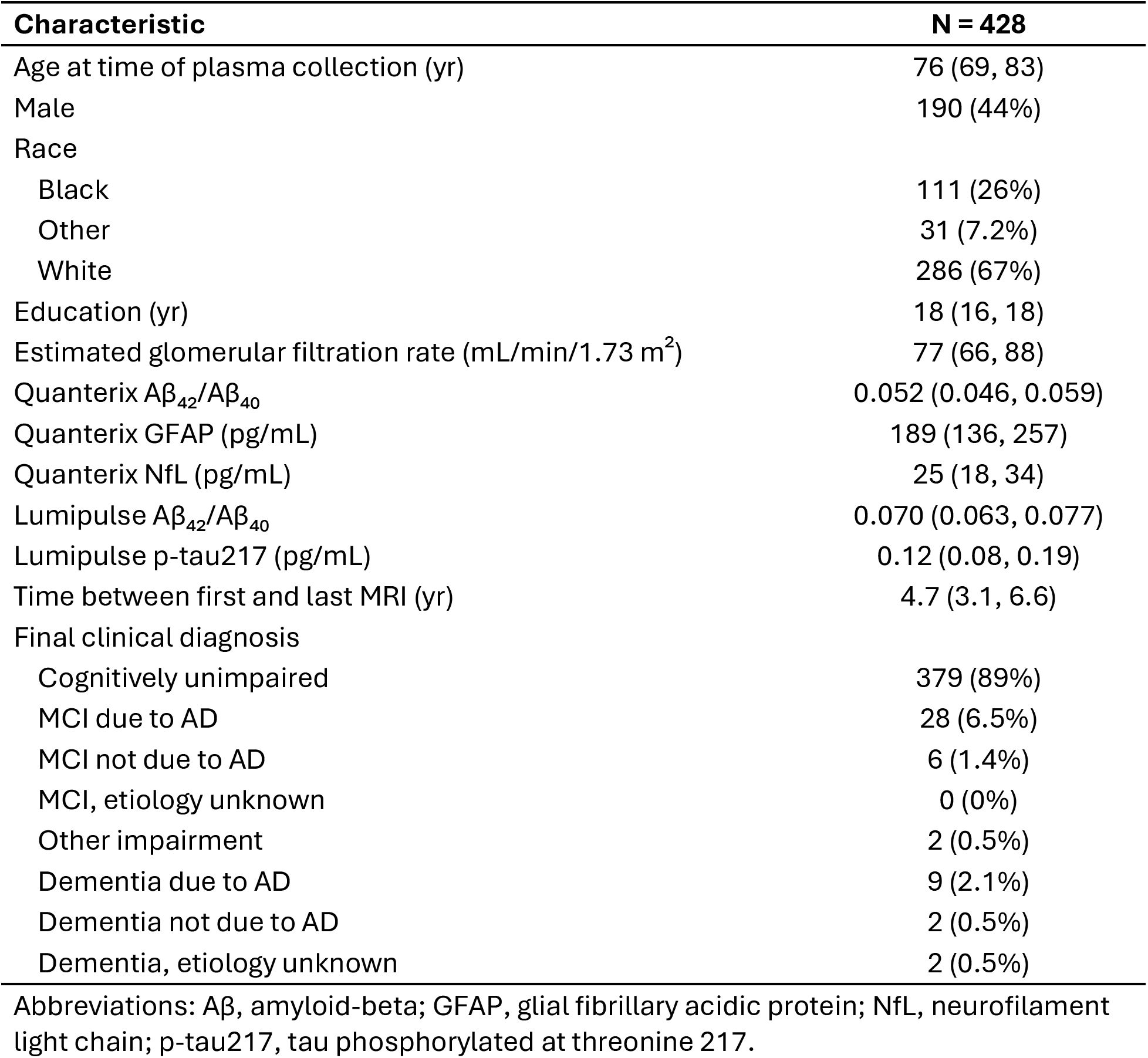
Participant characteristics for the Fujirebio Lumipulse sample. For continuous and categorical variables, we report the median and interquartile range or the N and percentage, respectively.

**Supplementary Figure 1.**
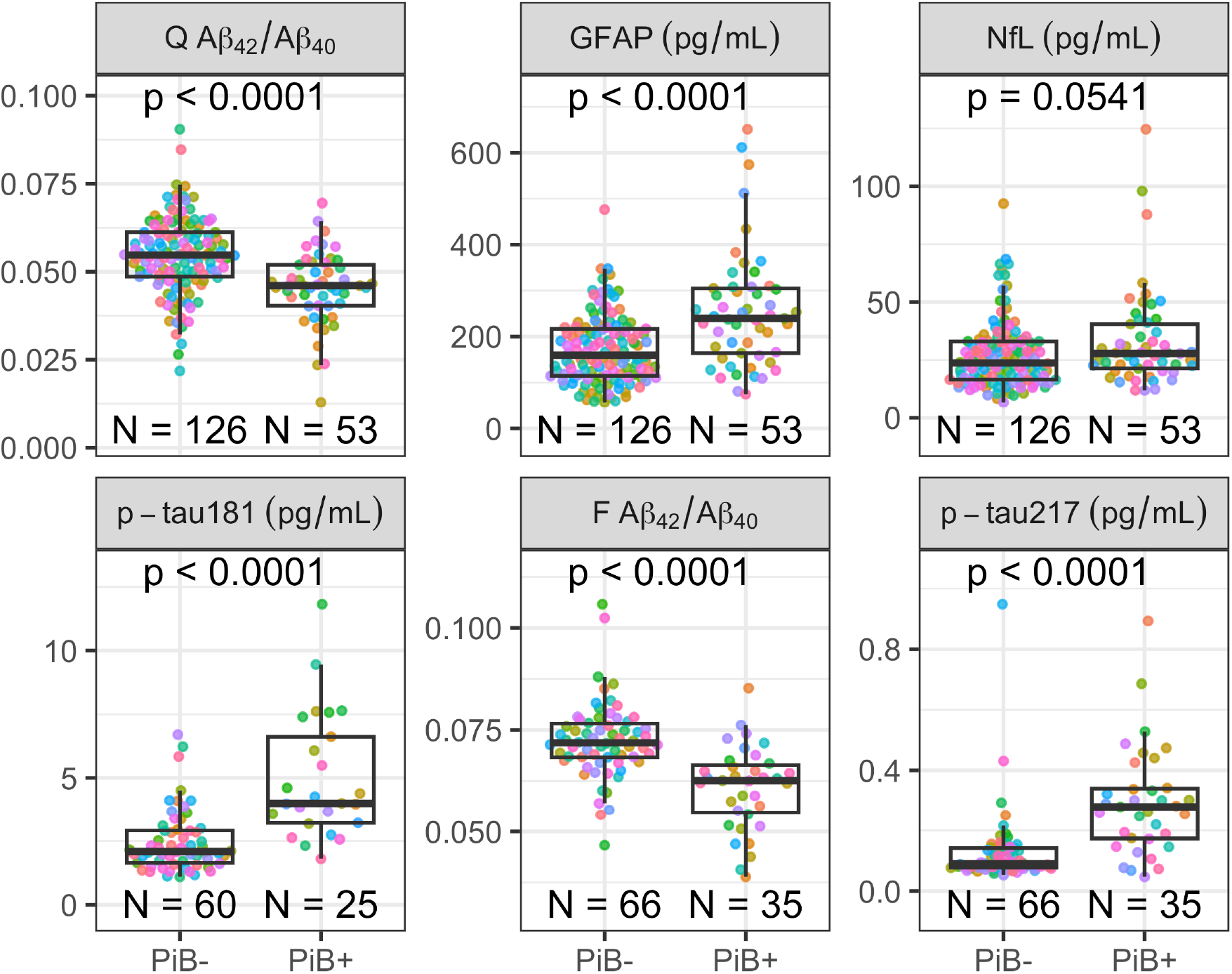
Plasma biomarkers at index visit by concurrent amyloid status as determined using [^11^C]PiB PET. All plasma biomarkers except for NfL exhibited statistically significant (p < 0.05) group differences as assessed with the Wilcoxon rank-sum test. Abbreviations: Aβ, amyloid-beta; F, Fujirebio Lumipulse; GFAP, glial fibrillary acidic protein; NfL, neurofilament light chain; p-tau181, tau phosphorylated at threonine 181; p-tau217, tau phosphorylated at threonine 217; PiB, Pittsburgh compound B; Q, Quanterix Simoa.

**Supplementary Table 3.** Cox proportional hazards model results for atrophy pattern scores. Table reports the associations of atrophy pattern scores (z-scored) at index visit with conversion to all-cause MCI/dementia (left column), MCI/dementia due to AD (middle), and due to non-AD (right).

| Atrophy score | All-cause MCI/Dem |  |  | Due to AD |  |  | Due to non-AD |  |  |
| --- | --- | --- | --- | --- | --- | --- | --- | --- | --- |
|  | HR | 95% CI | p | HR | 95% CI | p | HR | 95% CI | p |
| SPARE-AD | 1.84 | 1.47, 2.30 | <0.001 | 1.88 | 1.44, 2.46 | <0.001 | 1.52 | 0.97, 2.38 | 0.071 |
| SPARE-BA | 2.03 | 1.47, 2.81 | <0.001 | 1.65 | 1.14, 2.39 | 0.008 | 2.66 | 1.39, 5.11 | 0.003 |
| R1 | 1.07 | 0.87, 1.31 | 0.5 | 1.03 | 0.81, 1.32 | 0.8 | 1.12 | 0.73, 1.71 | 0.6 |
| R2 | 1.44 | 1.15, 1.80 | 0.001 | 1.50 | 1.15, 1.94 | 0.002 | 1.29 | 0.82, 2.03 | 0.3 |
| R3 | 1.30 | 1.02, 1.66 | 0.033 | 1.13 | 0.85, 1.50 | 0.4 | 1.73 | 1.05, 2.86 | 0.031 |
| R4 | 1.50 | 1.20, 1.88 | <0.001 | 1.16 | 0.89, 1.52 | 0.3 | 2.47 | 1.53, 3.98 | <0.001 |
| R5 | 1.56 | 1.23, 1.97 | <0.001 | 1.32 | 1.00, 1.74 | 0.047 | 1.98 | 1.22, 3.21 | 0.006 |

Abbreviations: AD, Alzheimer's disease; CI, confidence interval; HR, hazard ratio; MCI, mild cognitive impairment.

**Supplementary Table 4.** Cox proportional hazards model plasma biomarker results. Table reports the associations of plasma biomarkers at index visit (z-scored) with conversion to all-cause MCI/dementia (left column), MCI/dementia due to AD (middle), and due to non-AD (right).

| Biomarker | All-cause MCI/Dem |  |  | Due to AD |  |  | Due to non-AD |  |  |
| --- | --- | --- | --- | --- | --- | --- | --- | --- | --- |
|  | HR | 95% CI | p | HR | 95% CI | p | HR | 95% CI | p |
| QX A$\beta_{42}$/A$\beta_{40}$ | 0.82 | 0.67, 1.00 | 0.052 | 0.82 | 0.65, 1.04 | 0.11 | 0.80 | 0.52, 1.21 | 0.3 |
| QX log GFAP | 1.81 | 1.41, 2.33 | <0.001 | 1.46 | 1.07, 1.99 | 0.016 | 2.22 | 1.36, 3.61 | 0.001 |
| QX log NfL | 1.41 | 1.07, 1.86 | 0.016 | 1.22 | 0.87, 1.72 | 0.3 | 1.55 | 0.86, 2.80 | 0.15 |
| QX log p-tau181 | 1.61 | 1.20, 2.16 | 0.002 | 1.72 | 1.22, 2.44 | 0.002 | 1.16 | 0.63, 2.15 | 0.6 |
| FL A$\beta_{42}$/A$\beta_{40}$ | 0.62 | 0.46, 0.84 | 0.002 | 0.60 | 0.43, 0.84 | 0.003 | 0.82 | 0.37, 1.83 | 0.6 |
| FL log p-tau217 | 1.82 | 1.39, 2.39 | <0.001 | 1.79 | 1.30, 2.47 | <0.001 | 1.67 | 0.93, 3.00 | 0.083 |

Abbreviations: Aβ , amyloid-beta; AD, Alzheimer's disease; CI, confidence interval; FL, Fujirebio Lumipulse; GFAP, glial fibrillary acidic protein; HR, hazard ratio; MCI, mild cognitive impairment; NfL, neurofilament light chain; QX, Quanterix Simoa.

**Supplementary Figure 2.**
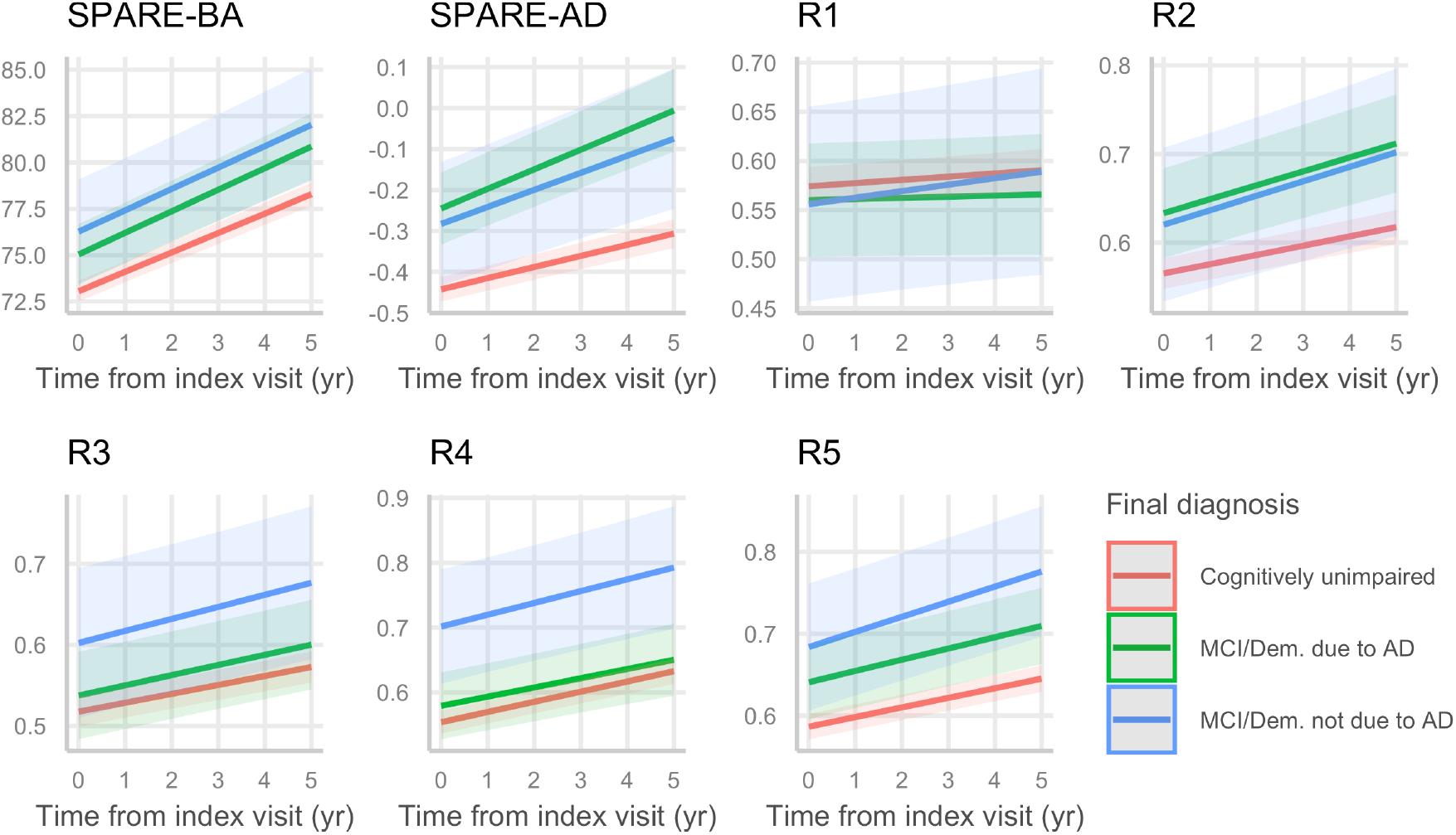
Longitudinal rate of change in atrophy scores by incident MCI/dementia. In demographics- and eGFR-adjusted models, compared to participants who remained cognitively unimpaired, those who converted to MCI/dementia due to AD had greater SPARE-BA, SPARE-AD, R2, and R5 at index visit, and exhibited steeper longitudinal increases in SPARE-AD and R2. Compared to participants who remained cognitively unimpaired, those who converted to MCI/dementia not due to AD had greater SPARE-BA, SPARE-AD, R4, and R5 at index visit, and exhibited steeper longitudinal increases in SPARE-AD and R5.

